# Multimodal Artificial Intelligence Fusion Models to Predict Response to Venetoclax and Azacitidine in Patients with Newly Diagnosed Acute Myeloid Leukemia

**DOI:** 10.64898/2026.10.12.26364938

**Authors:** Jan-Niklas Eckardt, Ishan Srivastava, Kellen Gil, Andrea Abeyta Osypuk, Zenggang Pan, Mohamed Benouis, Martin M. K. Schneider, Freya Schulze, Susann Winter, Katja Sockel, Martin Bornhäuser, Rabia Shahswar, Michael Heuser, Jan Moritz Middeke, Daniel A. Pollyea

## Abstract

Venetoclax and azacitidine (ven-aza) is a standard of care therapy for newly diagnosed acute myeloid leukemia (AML) patients unfit to receive intensive therapy, and may also be appropriate for certain fit patients. Models predicting response to ven-aza currently focus predominantly on genetics. Here, we present two multimodal fusion models: a custom ResNet-34-based model and a pathology foundation model fine-tuned for bone marrow smear (BMS) images that combine clinical and genetic variables with whole slide images. Training on 193 newly diagnosed AML patients who received ven-aza, we obtained an area-under-the-receiver-operating-characteristic (AUROC) of 0.79 and 0.82 for the ResNet-based and fine-tuned foundation models in a heldout test set, and 0.76 and 0.73 in an external validation set, predicting best response to this therapy. Sensitivity analysis showed that fusing image data with clinical and genetic variables outperformed training on either modality alone. In univariable and multivariable analyses, image-based classification was an independent predictive marker, second only in effect size to *IDH1/2* mutation status, outperforming other genetic alterations. Image-based explainability showed high model attention on myeloblasts, suggesting previously unrecognized cell-specific patterns associated with response. Our findings suggest BMS images complement genetic biomarkers to refine outcome prediction in ven-aza treatment and incentivize AI-supported outcome prediction, including bone marrow morphology.

## Introduction

The combination of the BCL-2-inhibitor venetoclax (Ven) with hypomethylating agents (HMA) such as azacitidine (Aza) is a therapeutic standard for newly diagnosed patients with acute myeloid leukemia (AML) who are unfit to receive intensive chemotherapy.^1^ Recent reports^2,3^ also suggest that patients eligible for intensive chemotherapy may benefit from Ven-Aza, reporting higher rates of complete remission (CR), fewer adverse events, and shorter durations of hospitalization. While a majority of patients respond, a significant minority do not^4,5^ and non-responders have particularly poor outcomes^6^. It would therefore be clinically useful to predict non-responders prior to the initiation of therapy, but predicting responses to Ven-Aza is a challenge. As a result, multiple recent studies investigating genetic biomarkers predictive of therapeutic sensitivity or resistance to ven-aza have been published.^7–10^ In general, alterations of *NPM1, IDH1,* and *IDH2* have been associated with improved response rates and survival, whereas alterations of *TP53*, *RAS*, and *FLT3*-ITD were reported to confer unfavorable outcomes.^11–17^ Yet, isolated mutations alone may not be sufficient to predict response. Multiple recent studies suggested phenotypic differentiation stage to affect outcomes to Ven-Aza, with patients with (myelo-)monocytic phenotypes (corresponding to the M4 and M5 subtype of the French-American-British classification^18^) reported to have lower rates of response. In addition, there appears to be direct interactions between morphology and certain gene mutations.^19–24^ For instance, monocytic AML, an adverse feature, and *NPM1* mutations, with positive prognostic significance, frequently co-occur and on these occasions, *NPM1* trumps monocytic morphology.^25^ Linking morphology to response assessment may provide a fast, affordable, and widely deployable screening method. However, a mere dichotomization of monocytic and non-monocytic phenotypes does not provide the predictive power to conclusively inform therapeutic decision making.

The capability of artificial neural networks to segment and classify image data, so-called computer vision, is currently deployed across multiple medical imaging modalities, including hematologic microscopy, where these algorithms have proven capable of correctly identifying cell types, classifying disease and predicting genotype-phenotype links.^26^ For instance, several recent studies report neural networks to directly predict genetic alterations in AML based on microscopic images from bone marrow or peripheral blood smears alone, including *NPM1*, *FLT3*-ITD, CBFB::MYH11 fusions or RUNX1::RUNX1T1 fusions, and t(15;17)/PML-RARa.^27–32^ Interestingly, medical images appear to hold an abundance of currently unused information^33^, potentially also informative of treatment outcomes. This has been demonstrated in histopathology images of solid cancers, where deep learning models analyze tissue composition and predict treatment responses and survival.^34^ For example, in colorectal cancer, a deep learning-based stroma score predicted patient survival^35^, in hepatocellular carcinoma, a deep learning model predicted outcomes after resection^36^, and in small cell lung cancer, a deep learning model identified phenotypic clusters associated with survival.^37^

In this study, we evaluated computer vision models to predict response to upfront ven-aza therapy at initial diagnosis using convolutional neural networks as well as state-of-the-art pathology foundation models, both on image data only and multimodally, incorporating image data and clinical/genetic features.

## Methods

### Data sets and preprocessing

Patients were eligible upon diagnosis of AML according to WHO^38^ or ICC^39^ definitions, age ≥ 18 years, upfront treatment with ven-aza, and available biomaterial including bone marrow smears (BMS) from initial diagnosis. Patients with acute promyelocytic leukemia (APL, AML-M3) were excluded. Whole-slide-images (WSI) of bone marrow smears from 193 newly diagnosed AML patients who received frontline treatment with ven-aza between June 2015 and December 2024 were digitized. This retrospective, de-identified analysis was deemed exempt from institutional review board (IRB) review by the Colorado multiple IRB. BMS were prepared and air-dried before staining with the Hema 3 staining system (Fisher Scientific, USA), a three-step staining comparable to the Wright–Giemsa method. WSI scanning was performed using the Leica Aperio AT2 automated microscope slide scanner at 40X magnification, corresponding to a spatial resolution of 0.25 µm per pixel. For karyotype analysis, standard techniques for chromosome banding and fluorescence-in-situ-hybridization were used. Molecular analysis was conducted using a proprietary diagnostic myeloid next generation sequencing panel with a variant allele fraction cut-off of 5% binarizing molecular alterations. External validation data was obtained from Hannover Medical School, Hannover, Germany, comprising 19 patients with newly diagnosed AML and treated with ven-aza. Diagnostic mutation profiling of the external data set was performed by targeted sequencing of recurrently mutated AML genes using a custom myeloid panel sequenced on the MiSeq platform as described previously.^40^ Genomic positions with a read depth of less than 100 reads and detected variants with a VAF <5% were excluded. *ASXL1* c.1934 dupG was excluded if the VAF was <18%. These samples were scanned using the Pannoramic SCAN II (3DHISTECH, Budapest, Hungary) equipped with a 40x objective and a 0.63x C-mount adapter, yielding a spatial resolution of 0.1377 µm per pixel. All patients were retrospectively categorized according to the ELN 2024 Less-Intensive^7^ risk stratification system. Karyotypes were additionally categorized according to the Southwest Oncology Group/Eastern Cooperative Oncology Group Study system.^41^ Response was defined as complete remission (CR) or complete remission with incomplete hematologic recovery (CRi) in accordance with ELN 2022 recommendations.^42^ Any state less than CR/CRi was defined as a non-response.

### Computer Vision Models

We employed multiple evaluation strategies in this study. First, we developed a custom multi-instance learning model using BMS images only, incorporating a patch-based BMS image filtering method. Second, state-of-the-art transformer-based^43^ pathology vision models were fine-tuned for BMS-based response prediction. Third, we implemented a late multimodal fusion strategy, adding clinical and genetic information to BMS image data for both our custom model and the fine-tuned pathology foundation models. Figure 1 displays the three-tiered study workflow.

**Figure 1.**
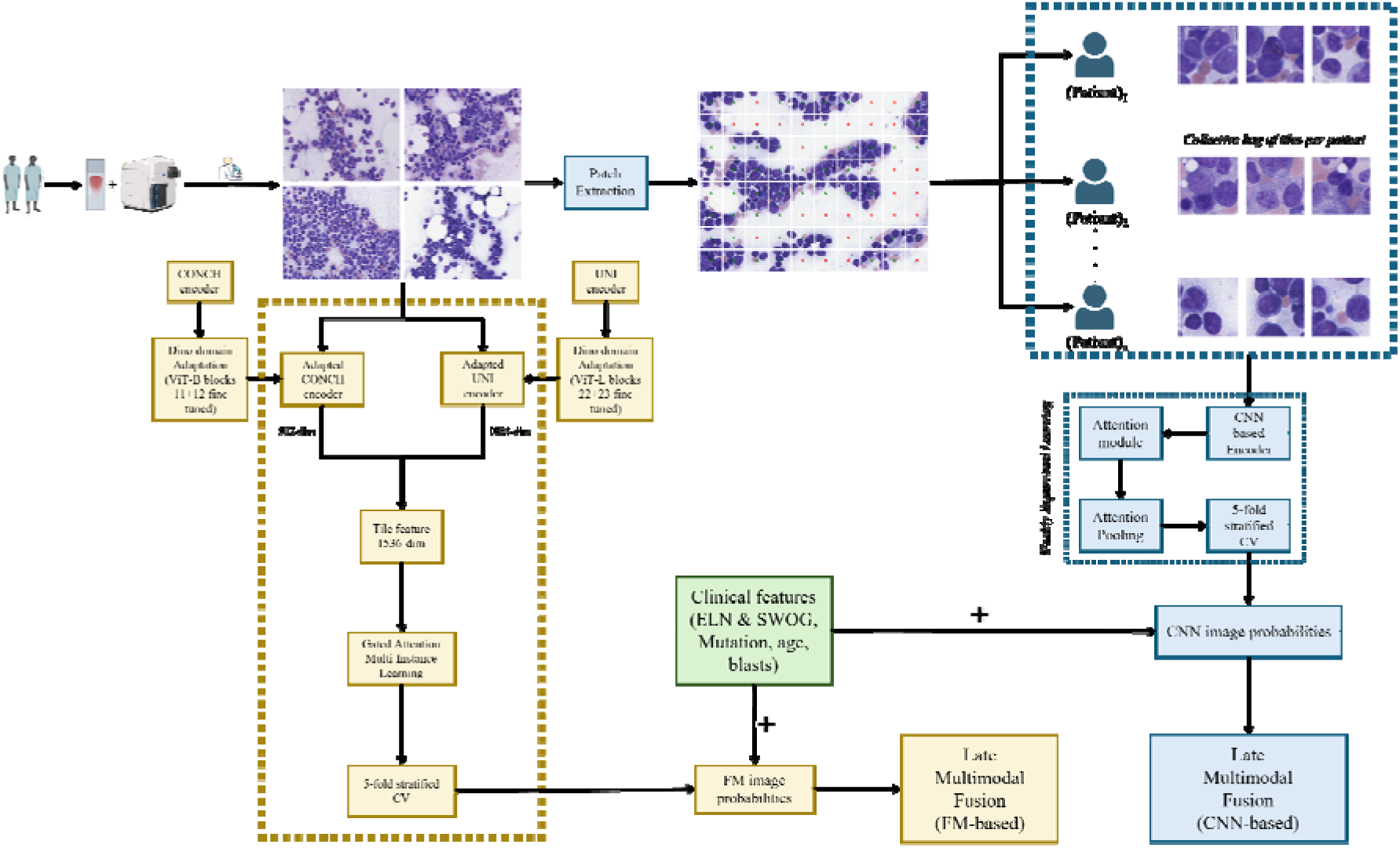
End-to-end computational pipeline for venetoclax/azacitidine treatment response prediction from bone marrow smears. Digitized bone marrow smears were evaluated by two independent pipelines: a foundation model (FM) approach using concatenated CONCH and UNI pathology foundation models as well as a fine-tuned convolutional neural net (CNN; ResNet-34) to generate patient-level image probabilities. Both pathways parallely combine image probabilities with clinical and genetic features in a late multimodal fusion step.

### Patch extraction and adaptive quality control

For our custom model, WSI were manually segmented for regions-of-interest (ROI), evaluating diagnostically relevant and appropriate cellular content in areas between bone marrow spicules, sharp image focus, staining quality, and absence of image artifacts, thereby excluding regions with only peripheral blood, poor image or staining quality, or areas scanned out of focus. ROIs were further partitioned into 358*358 pixel, non-overlapping, non-padded patches by cropping each ROI to the nearest integer multiple of the patch size, discarding at most 50 pixels along each border dimension. To account for variability in image quality between slides (considering differences in stain quality and specimen age), we implemented a region-adaptive quality control (QC) mechanism that dynamically adjusts filtering thresholds based on the statistical properties of each image rather than being fixed globally. For each ROI, mean and standard deviation of hue, saturation and luminance were computed, allowing filtering thresholds to adapt to ROI-specific color distributions, mitigating potential effects of global staining bias. Next, we implemented a patch-based filtering step to exclude patches predominantly consisting of cell-free background or erythrocyte-dominant regions (Figure 2), minimizing the number of non-informative patches without nucleated cells for downstream custom model development.

**Figure 2.**
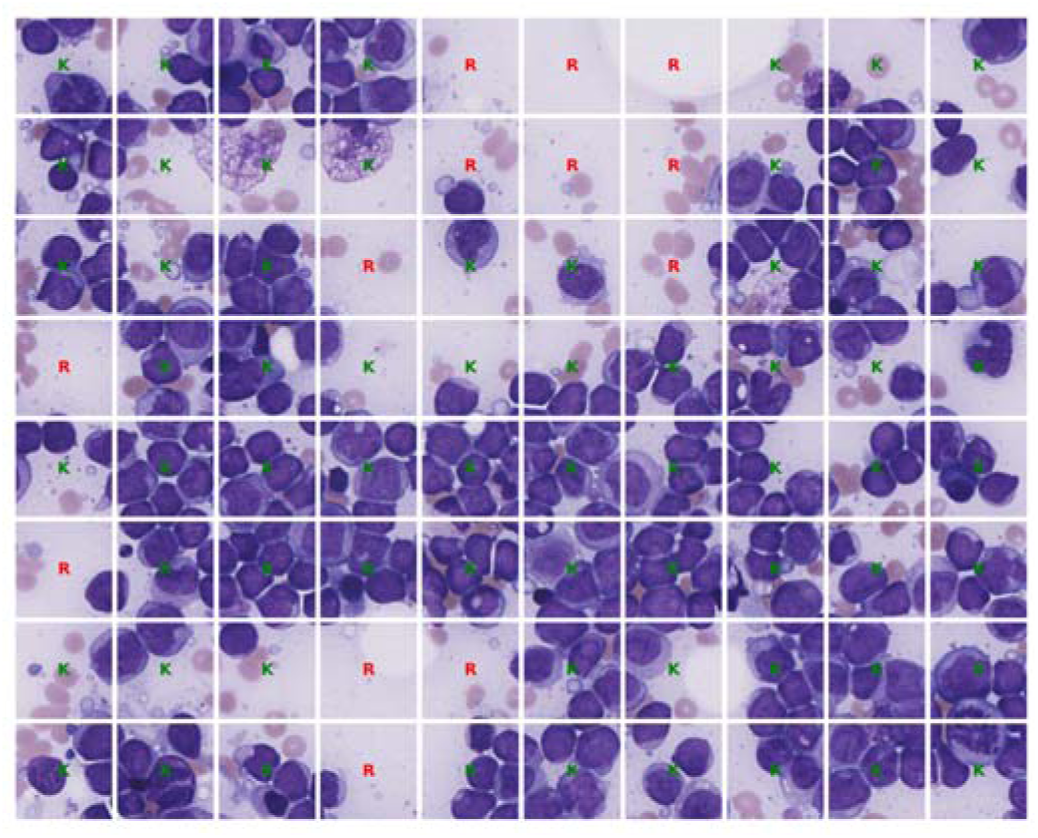
Representative example of patch extraction and adaptive quality control. The image was divided into a 358*358 pixel grid, and the quality control step filters these tiles into two categories: Kept (marked as K in green) and Rejected (marked as R in red) based on cellularity.

### Multi-Instance Learning

In our custom model, patch-level features were first extracted for each patient separately using a convolutional encoder (ResNet-34^44^) and were then passed through a small neural attention module, consisting of two fully connected layers with a non-linear activation function (sigmoid and tanh), thereby producing a scalar attention score for each tile. These scores were normalised using a softmax function to yield a distribution of attention weights, allowing the model to learn the relative importance of different patches within the cohort. The softmax was computed over all quality controlled patches, so that every patch contributed in proportion to its learned weight. The final patient representation was then obtained by computing a weighted average of patch features, where weights were determined by the refined attention scores. This aggregated feature vector was then passed through a fully connected classification head, followed by a sigmoid activation function to produce a single prediction per patient which was interpreted as a probability. Finally, the entire model (feature extractor, attention module, and classifier) was trained end to end using binary cross entropy loss at the patient level. Training was conducted using the Adam optimizer^45^ with momentum, learning rate scheduling, and gradient clipping. Mixed precision training^46^ was used to optimize memory usage for the larger patch sets (104,378 patches).

### Foundation Models

Image-level feature representations were extracted using two state-of-the-art, pathology-specific foundation models: Contrastive Learning from captions for histopathology (CONCH)^47^ and Universal model of Pathology (UNI)^48^. CONCH is a vision language model pretrained on over 1.17 million pathology image-caption pairs using contrastive learning, and UNI is a pathology encoder pretrained via self-supervised learning on 100,000 whole slide images spanning 20 tissue types. Importantly, neither UNI nor CONCH have been trained on BMS image data. To adapt the CONCH and UNI encoder to the specific morphological characteristics of bone marrow slides, we performed a self-supervised domain adaptation using the self-distillation with no labels (DINO)^49^ framework. The final two transformer blocks (10 and 11) of the CONCH ViT-B/16 backbone as well as the corresponding blocks (22+23) of the UNI ViT-L backbone were fine tuned on all the bone marrow images using a student-teacher distillation objective without any response labels, while the remaining encoder weights were kept frozen. To further improve the generalizability of the extracted features, embeddings from the adapted CONCH and UNI models were concatenated along the feature dimension to produce a 1536-dimensional combined representation. Concatenation was preferred over selecting a single encoder because of their complementary pre-trained information: CONCH captures semantic pathology, while UNI understands morphological textures.

### Late Multimodal Fusion

Following image-only outcome prediction, we hypothesized that, since genetic alterations and clinical variables may affect response to ven-aza, adding these variables to images in a multimodal fusion approach could potentially boost predictive performance. Hence, for both our custom MIL model and the fine-tuned foundation models, a two-stage late multimodal fusion was applied. In the first stage, image-only response probabilities were generated for all 193 patients through five-fold cross-validation by both vision model pipelines. In the second stage, logistic regression (LR) with L2 regularization, random forest, elasticnet and XGBoost were trained to predict response from a 17-dimensional feature vector, consisting of image probability and 16 clinical/genetic features: ELN 2024 less intensive risk (favorable, intermediate, adverse), SWOG cytogenetic category (favorable, intermediate, adverse), binarized mutation status for *NPM1*, *IDH1*/2, *TP53*, *ASXL1*, *RAS*, *PTPN11*, *RUNX1* as well as *FLT3*-ITD, age at diagnosis (normalised to [0,1]) and baseline bone marrow blast percentage (normalised to [0,1]). The final fusion models were trained and evaluated using the same five-fold cross-validation data distribution as the image-only models, ensuring that these models cannot access the response labels of the held out patients. Lastly, to account for the stochastic variability inherent to neural network training on small cohorts, all experiments were repeated with three independent random seeds governing weight initialisation, data augmentation sampling and fold shuffling. Patients whose cytogenetic risk could not be assigned were represented by the all-zero indicator vector, i.e. treated as the reference level rather than dropped or imputed.

### Performance evaluation

Model performance was evaluated using accuracy, average precision (AP) and receiver operating characteristic area under the curve (ROC-AUC) for prediction of ground truth labels (responder vs. non-responder), where each patient is represented as a bag of image tiles and assigned a single ground-truth label. Given the class imbalance of responders and non-responders, prediction thresholds for each model were selected according to their optimal Youden threshold^50^. Validation was performed on a heldout test set with five-fold stratified cross-validation. Confidence intervals were computed via 2000 bootstrap resamples with replacement from the pooled out-of-fold predictions at the Youden-optimal threshold.

### Qualitative and quantitative model interpretability

Image areas predictive of response or non-response were analyzed according to model attention at two levels. Computer vision model attention for the foundation branch was visualized using attention rollout.^51^ Rollout is computed on each 224*224 pixel tile, which is represented by a ViT as 196 patch token (block of 14*14 grid at 16-pixel resolution) together with a class token. These blocks repeatedly interact and exchange information with each other and we trace this exchange to its advent, forming the basis of a relevance score. This score depicts which block was responsible, to what degree, in forming the representation of that particular tile, thus providing us with a rollout heatmap, enabling human-in-the-loop morphological evaluation of these image areas. Quantitatively, feature attributions were computed with SHAP (SHapely Additive exPlanations). Within each cross-validation fold, a linear SHAP explainer with correlation-dependent perturbation was applied to the held-out fold. Since the folds partition the cohort, every patient was explained exactly once by a model iteration that had not seen them, yielding one attribution vector per patient in log-odds of response. The reported baseline is the mean expected value across the five fold explainers.

## Data availability

Image data used for training and testing computer vision models are available upon reasonable request from the corresponding author.

## Code availability

The code generated for the purpose of this project is available <u>here</u>. https://github.com/Ai-in-Cancer-org/venAza

## Results

### Computer vision models accurately predict response to venetoclax azacitidine on bone marrow smear images alone

193 newly diagnosed adult AML patients who received firstline therapy with ven-aza were included in this study. Median age was 70 years (interquartile range [IQR]: 65-76) and 42.0% of patients were female. Median bone marrow blast count at initial diagnosis was 50% (IQR: 30-77%). 63.2% of patients had ELN2024 less-intensive favorable risk, 25.9% had intermediate risk, and 10.9% had adverse risk. CR/CRi as best response was achieved in 79.8%, with a median treatment duration until best response of 35 days (IQR: 30-49). Table S1 provides detailed baseline characteristics.

First, we trained a custom ResNet-34-based model with adaptive tile filtering and MIL to predict response status directly from BMS images without incorporating clinical or genetic variables. In five-fold stratified cross-validation, the image-only neural network classifier achieved a pooled out-of-fold (OOF) AUROC of 0.753 (95%-CI: [0.678-0.829], Figure 3A), with pooled accuracy of 0.819 (95%-CI: [0.758-0.867]), F1 score of 0.887 (95%-CI: [0.848, 0.922]), sensitivity of 0.890 (95%-CI: [0.830-0.930]), specificity of 0.538 (95%-CI: [0.386-0.685]), PPV of 0.884 (95%-CI: [0.829, 0.933]), and NPV of 0.553 (95%-CI: [0.395, 0.707]). Full per-fold metrics are provided in Table S2.

**Figure 3.**
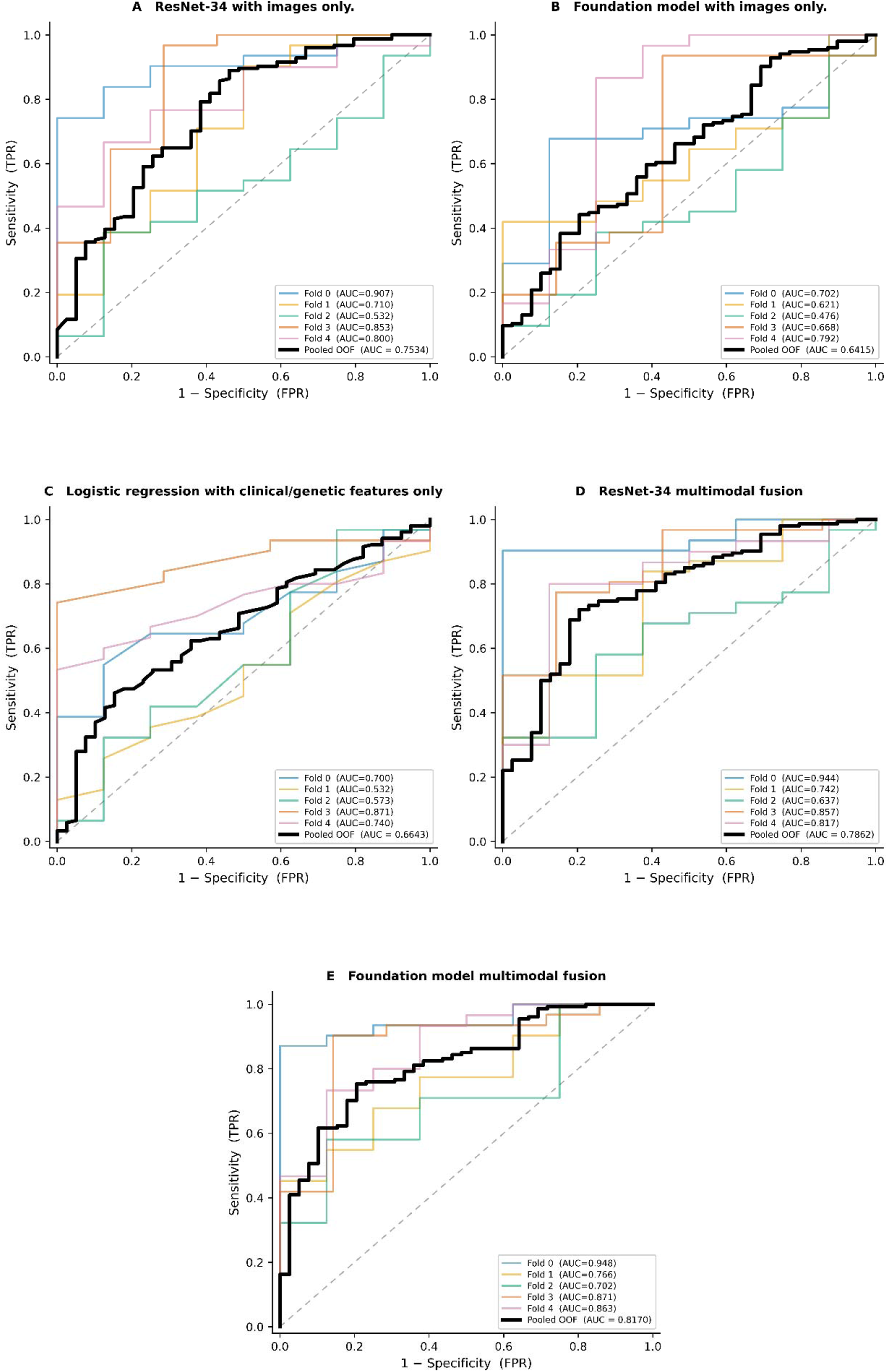
Receiver operating characteristic (ROC) curves for the fusion and non-fusion models across 5-fold cross-validation. (A) Image-only custom model (ResNet-34). (B) Image-only foundation model (CONCH+UNI). (C) Clinical/genetic feature-only logistic regression-based classifier. (D) ResNet-34-based fusion model using logistic regression. (E) Foundation model fusion using logistic regression. Individual fold curves are shown in colour; the bold black line represents the pooled out-of-fold (OOF) estimate.

Next, we fine-tuned the self-supervised pathology foundation models UNI and CONCH on the same task using the same data distribution to maintain comparability. Using BMS images only, the foundation model achieved a pooled OOF AUROC of 0.641 (95%-CI: [0.541, 0.738], Figure 3B), with pooled accuracy of 0.513 (95%-CI: [0.440, 0.580]), F1 score of 0.591 (95%-CI: [0.514, 0.661]), sensitivity of 0.442 (95%-CI: [0.364, 0.519]), specificity of 0.795 (95%-CI: [0.660, 0.917]), PPV of 0.895 (95%-CI: [0.821, 0.961]), and NPV of 0.265 (95%-CI: [0.186, 0.345]). Full per-fold metrics are provided in Table S3.

### Late multimodal fusion of bone marrow smear images with clinical and genetic patient features improves response prediction to venetoclax-azacitidine

Previous reports indicated that response to ven-aza is associated with genetic alterations. As a baseline, we used patient information on ELN 2024 Less-intensive risk category (favorable, intermediate, adverse), SWOG cytogenetic category (favorable, intermediate, adverse), binarized mutation status for *NPM1*, *IDH1*/2, *TP53*, *ASXL1*, *RAS*, *PTPN11, RUNX1* as well as *FLT3*-ITD, age at diagnosis, and baseline bone marrow blast percentage for response prediction via four non-image-based classifiers (logistic regression with L2 regularization, random forest, elasticnet and XGBoost). Logistic regression outperformed all other classifiers, achieving a pooled OOF AUROC of 0.6643 (95%-CI: [0.576, 0.752], Figure 3C), an accuracy of 0.544 (95%-CI: [0.474, 0.613]), F1 score of 0.619 (95%-CI: [0.520–0.672]), sensitivity of 0.461 (95%-CI: [0.384, 0.540]), specificity of 0.846 (95%-CI: [0.703, 0.927]), PPV of 0.922 (95%-CI: [0.840–0.964]), and NPV of 0.284 (95%-CI:[ 0.210–0.372]), using non-image data only. Classification performance for the best performing classifier, i.e., logistic regression, is provided in supplementary Table S4.

Subsequently, we added clinical and genetic information in late multimodal fusion to BMS image data for both our custom ResNet-34-based model with adaptive filtering and the pathology foundation model. Using the fine-tuned foundation model with late multimodal fusion, we obtained a pooled OOF AUROC of 0.817 (95%-CI: [0.745, 0.884], Figure 3E), F1 score of 0.835 (95%-CI: [0.787, 0.881]), sensitivity of 0.753 (95%-CI: [0.684, 0.819]), specificity of 0.795 (95%-CI: [0.667, 0.914]), PPV of 0.935 (95%-CI: [0.889, 0.975]), and NPV of 0.449 (95%-CI: [0.333, 0.567]). For our custom model, late multimodal fusion of image and clinical/genetic patient data yielded a pooled OOF AUROC of 0.786 (95%-CI: [0.707, 0.862], Figure 3D), with pooled accuracy of 0.736 (95%-CI: [0.674, 0.793]), F1 score of 0.813 (95%-CI: [0.762, 0.859]), sensitivity of 0.721 (95%-CI: [0.651, 0.798]), specificity of 0.795 (95%-CI: [0.667, 0.923]), PPV of 0.933 (95%-CI: [0.885, 0.975]), and NPV of 0.419 (95%-CI: [0.304, 0.532]). Corresponding detailed tables for foundation fusion and CNN fusion models are provided in supplementary Tables S5-S8 and S9, respectively. Class-wise patient distributions for the fusion models are presented as confusion matrices in supplementary Table S10.

### Image-based response prediction is an independent marker of response to ven-aza

Univariable regression and Shapley additive explanations (SHAP) were used to quantify effect sizes of individual features in the multimodal models. In univariable regression, *IDH1/2* mutation status had the highest OR for CR prediction (OR 16.34, *p*=0.007), followed by foundation model image classification probability (OR 10.45, *p*<0.001), ResNet image classification probability (OR 8.81, *p*<0.001), and mutation status of *NPM1* (OR 6.45, *p*=0.012). In contrast, mutation status of *RAS* was associated with refractory disease (OR 0.25, *p*=0.003; Table 1). In a multivariable model adjusted for *NPM1*, *IDH1/2* and *RAS* mutation status, both foundation model image classification probability (multivariable OR 4.97 95%-CI 1.96-12.58, *p*=0.001) and ResNet image classification probability (multivariable OR 3.85 95%-CI 1.53-9.68, *p*=0.004) maintained their independent prognostic impact. To compare individual fusion model parameters, we calculated odds ratios for CR as best response additionally for ELN2024 less-intensive risk categories (although ELN2024 less-intensive categories were not used as a variable during model training) by offsetting intermediate risk as a reference (at OR=1) due to multicollinearity between the mutually exclusive categories. Patients with ELN2024 favorable risk had significantly higher odds to achieve CR (univariable OR 3.57 95%-CI 1.73-7.33, p=0.001), while odds for patients with ELN2024 adverse risk showed no statistical significance (univariable OR 1.00 95%-CI 0.34-2.93, *p*=0.998).

**Table 1.**
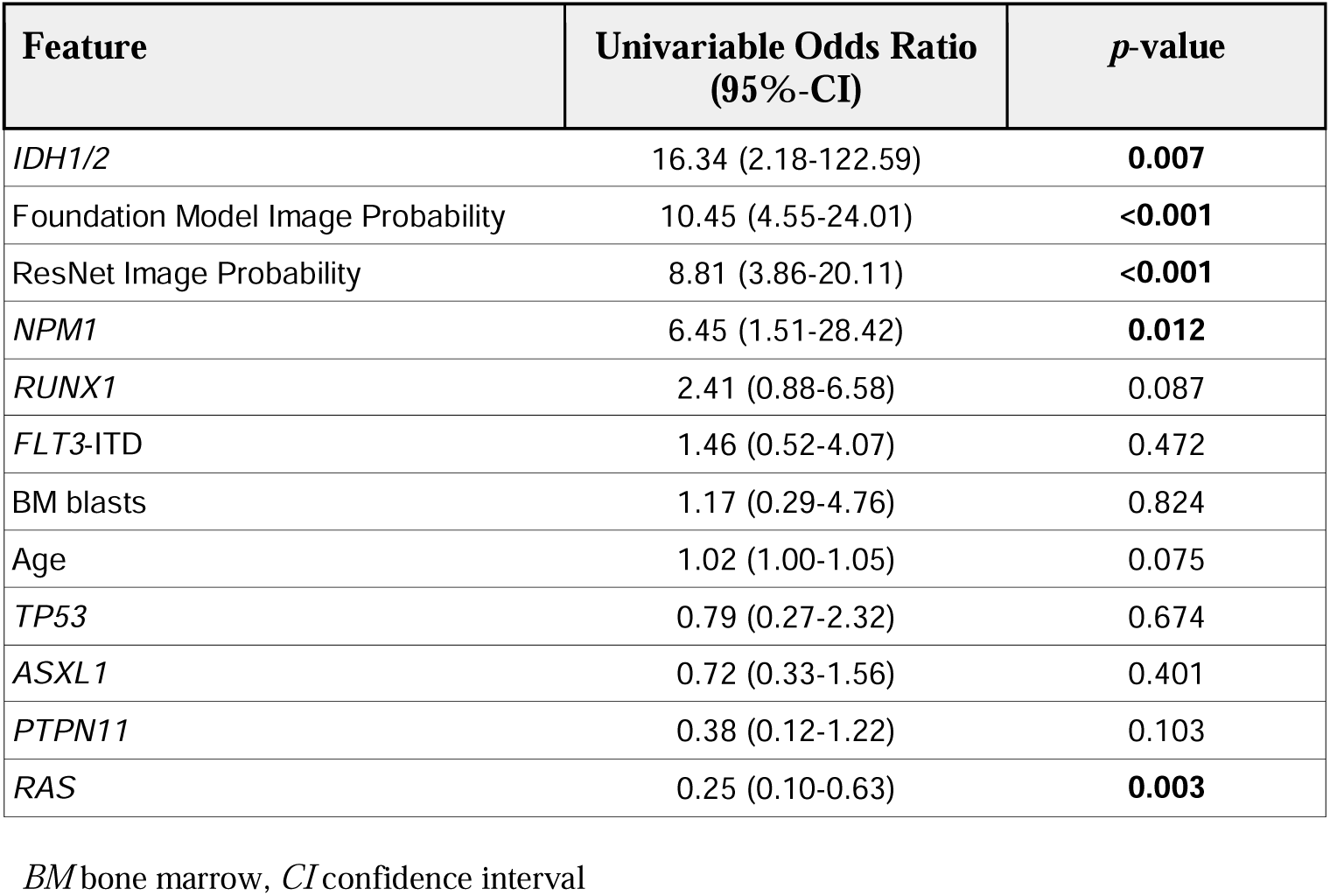
Univariable odds ratios for response prediction to venetoclax azacitidine. We found patients with *IDH1/2* mutations to have the highest odds to respond (CR/CRi) to ven-aza, closely followed by image-based response prediction with either the pathology foundation models or our custom ResNet-based model. Boldface indicates statistical significance (*p*<0.05).

### Quantitative and qualitative model explainability

In accordance, SHAP analysis showed that both fusion models (foundation model and ResNet-based custom model) were strongly impacted by alterations of *IDH1/2*, SWOG cytogenetics, image-based response prediction as well as alterations of *NPM1*, *RAS,* and *RUNX1*, pushing the decision boundary towards response prediction in true responders as well as towards prediction of refractory disease in true non-responders (Fig. 4). Beeswarm plots displaying SHAP values of each prognostic variable per individual patient (Fig. 4A, C) showed a scattered pattern, indicating that presence or absence of any given variable did not uniformly predict outcome across patients, but effect size rather varied between patients. On average, the largest SHAP effect sizes were measured for SWOG intermediate cytogenetics (+0.381 for response prediction and -0.675 for prediction of refractory disease) followed by *IDH1/2* mutation status, SWOG unfavorable cytogenetics, and image-based response probabilities ranking fourth (+0.131 for response and -0.189 for refractory disease) for the foundation models (Fig. 4B). Using our custom CNN model, the largest effect size again was measured for SWOG intermediate cytogenetics, followed by *IDH1/2* mutation status, SWOG unfavorable cytogenetics, *NPM1* alterations, and, either SWOG favorable cytogenetics and image-based probability in predicting response, or *RUNX1* alterations and image-based probability in predicting refractory disease (Fig. 4D).

**Figure 4.**
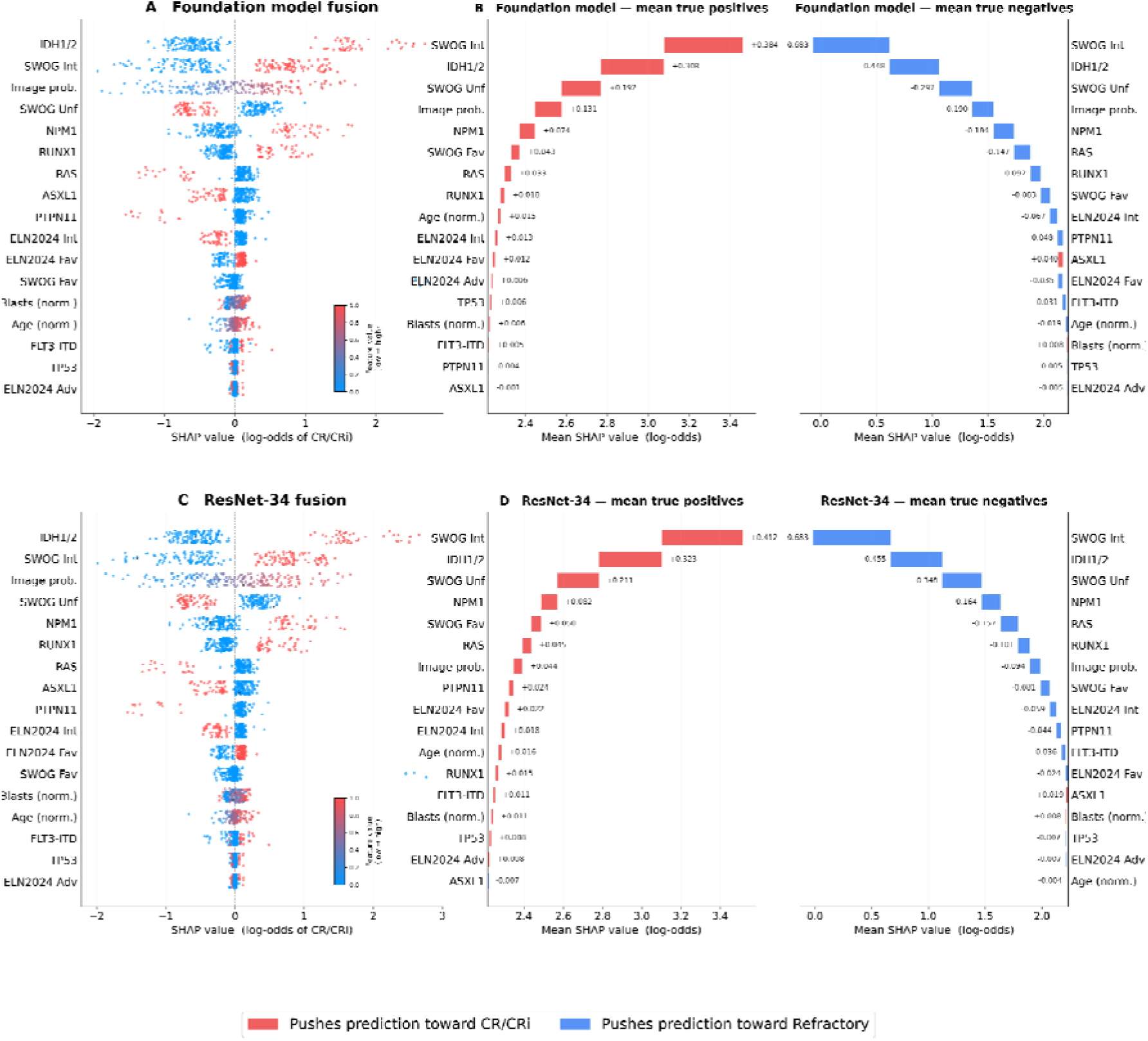
SHAP-based feature importance analysis for multimodal fusion models. SHAP values for the foundation model and the ResNet-34 model with logistic regression-based late fusion are reported on a log-odds scale. (A and C) *Beeswarm plots*: Each dot represents a patient, with the x-axis showing the SHAP values, i.e., the contribution of each feature towards the models’ predictions. Positive values push towards response, whereas negative values push towards refractoriness. (B and D) *Waterfall plots:* Mean SHAP values for the foundation and ResNet-34 fusion models, respectively, showing the average feature contributions for correctly predicted responders (true positives) and refractory patients (true negatives) on the log-odds scale.

Patients predicted to respond to ven-aza by the foundation fusion model had significantly higher rates of mutated *NPM1* (32.8% of true positives, 0% of true negatives, 25% of false positives, 5.4% of false negatives; Fisher’s exact *p*<0.001), mutated *IDH1/2* (39.7% of true positives, 0% of true negatives, 12.5% of false positives, 0% of false negatives; *p*<0.001), and, with borderline statistical significance, mutated *RUNX1* (26.7% of true positives, 6.5% of true negatives, 37.5% of false positives, 24.3% of false negatives; *p*=0.05). In contrast, mutations of *RAS* were significantly more frequent in patients correctly predicted not to respond (40.9% of true negatives, 6.9% of true positives, 12.1% of false negatives, 12.5% of false positives; *p*=0.01). There was no difference between TP, TN, FP and FN for mutation status of *TP53* (*p*=0.158), *ASXL1* (*p*=0.446), *PTPN11* (*p*=0.273), or *FLT3*-ITD (*p*=0.166).

Patients correctly predicted by the foundation model to respond had significantly higher bone marrow blast counts (true positive median 57%, IQR 35-80%) than patients correctly predicted not to respond (true negatives median 45%, IQR 35-83%) or patients with false positive (median 34%, IQR 23-86%) or false negative predictions (median 30%, IQR 23-66%; Kruskal Wallis *p*=0.0095). While patients correctly predicted by the foundation model to respond were statistically significantly older (true positives median age 72 years, IQR 67-77; false positives median age 71 years, IQR 64-76) compared to patients predicted to be refractory (true negatives median age 68 years, IQR 63-72; false negatives median age 69 years, IQR 62-76; Kruskal Wallis *p*=0.0403), the clinical significance of this age difference was negligible. There was no significant difference in patient sex between TP, TN, FP and FN (*p*=0.114).

In concordance analysis, both fusion models showed high agreement in predicting response (Pearson r=0.948, *p*<0.001) and refractory disease (Pearson r=0.913, *p*<0.001). In discordant predictions of response, the fusion foundation model correctly identified nine responding patients that were misclassified by the ResNet-based fusion model, whereas the latter correctly identified four responding patients that were misclassified by the fusion foundation model (Fig. 5A). Notably, among the nine responders uniquely identified by the fusion foundation model, five harbored *TP53* alterations. In contrast, discordant predictions of refractory disease were balanced between models, with each model correctly identifying two refractory cases that were misclassified by the respective other model (Fig. 5B). All discordantly predicted cases are summarized in supplementary Table S11.

**Figure 5.**
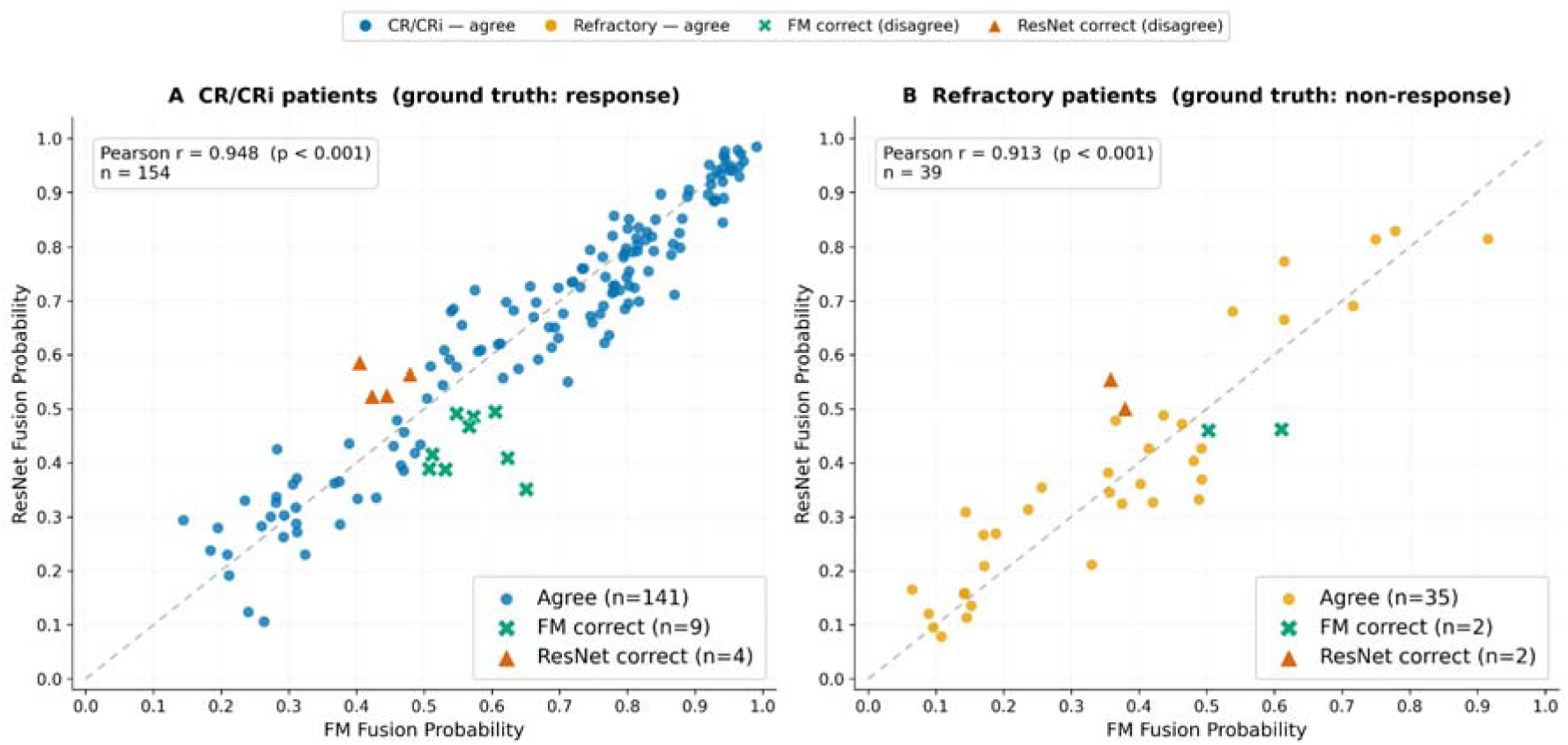
Scatter plot of foundation model (FM) versus ResNet-34 fusion probabilities for individual patients, stratified by ground-truth response status. (A) CR/CRi patients (n = 154). (B) Refractory patients (n = 39). Each point represents one patient. Filled (◘) circles indicate cases where both models agree at corresponding Youden’s threshold. Crosses (✕) indicate disagreement cases where the fusion FM predicted correctly. Triangles (▴) indicate disagreement cases where the ResNet-34-based fusion model predicted correctly. The dashed diagonal line represents perfect agreement between the two models. Pearson correlation coefficients and the total number of disagreements per panel are shown.

Image-based qualitative analysis using transformer rollout showed high cell-specific model attention, rather than focusing on background or artefacts, both for true positive and true negative predictions. In true positives (Fig. 6A-B), transformer attention was commonly placed on image areas containing myeloblasts, while attention on other cells was sparse. In true negatives (Fig. 6C-D), myeloblasts also received high attention, yet some attention was also diverted to other cell classes including lytic cells or smudges.

**Figure 6.**
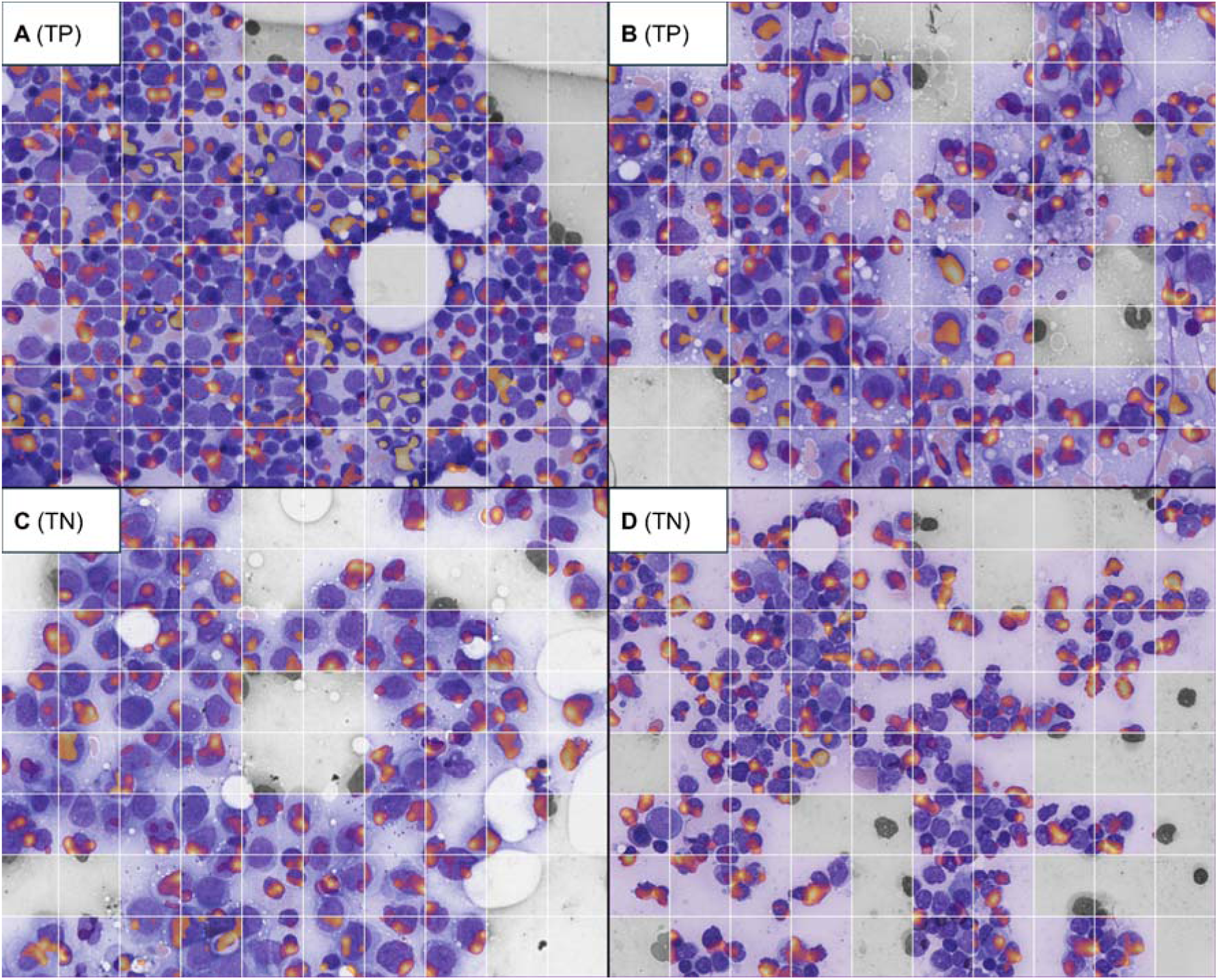
**Attention rollout indicates cell-specific model attention**, with a common focus on myeloblasts in both correctly predicted responders (true positives, TP; panels A and B) and correctly predicted non-responders (true negatives, TN; panels C and D). In TP, attention on other cells was rare, whereas in TN, some model attention was diverted towards other cell classes including lytic cells or smudges. Grey areas are tiles that were blocked after filtering and pre-processing due to sparse cell density.

### External validation

An independent external cohort of 19 newly diagnosed AML patients (13 responders and 6 non-responders) treated with ven-aza were obtained from Hannover Medical School, Germany. Four to six ROIs were manually selected per WSI, depending on image quality and cellularity. Macenko normalisation was used to minimize the impact of domain shifts in image properties, given the diverging staining patterns and camera properties between the original training/testing cohort and the external validation cohort. Our custom ResNet-34-based model using XGBoost for late multimodal fusion achieved an AUROC of 0.782 (95%-CI: [0.54-0.97]; Fig. 7A), with an accuracy of 0.737 (95%-CI: [0.51-0.88]), F1 score of 0.828 (95%-CI: [0.64-0.95]), sensitivity of 0.923 (95%-CI: [0.67–0.99]), specificity of 0.333 (95%-CI: [0.10–0.70]), PPV of 0.750 (95%-CI: [0.51-0.90]), and NPV of 0.667 (95%-CI: [0.21-0.94] in external validation. Detailed performance metrics are summarised in supplementary Table S12. The fine-tuned foundation model with late multimodal fusion using XGBoost achieved an AUROC of 0.718 (95%-CI: [0.36-1.00]; Fig. 7B), with an accuracy of 0.684 (95%-CI: [0.46-0.85]), F1 score of 0.769 (95%-CI: [0.55-0.92]), sensitivity of 0.769 (95%-CI: [0.50-0.92]), specificity of 0.500 (95%-CI: [0.19-0.81]), PPV of 0.769 (95%-CI: [0.50-0.92]), and NPV of 0.500 (95%-CI: [0.19-0.81]). Detailed performance metrics are summarised in supplementary Table S13. Class-wise patient distributions for the fusion models across the external cohort are presented as confusion matrices in supplementary Table S14.

**Figure 7.**
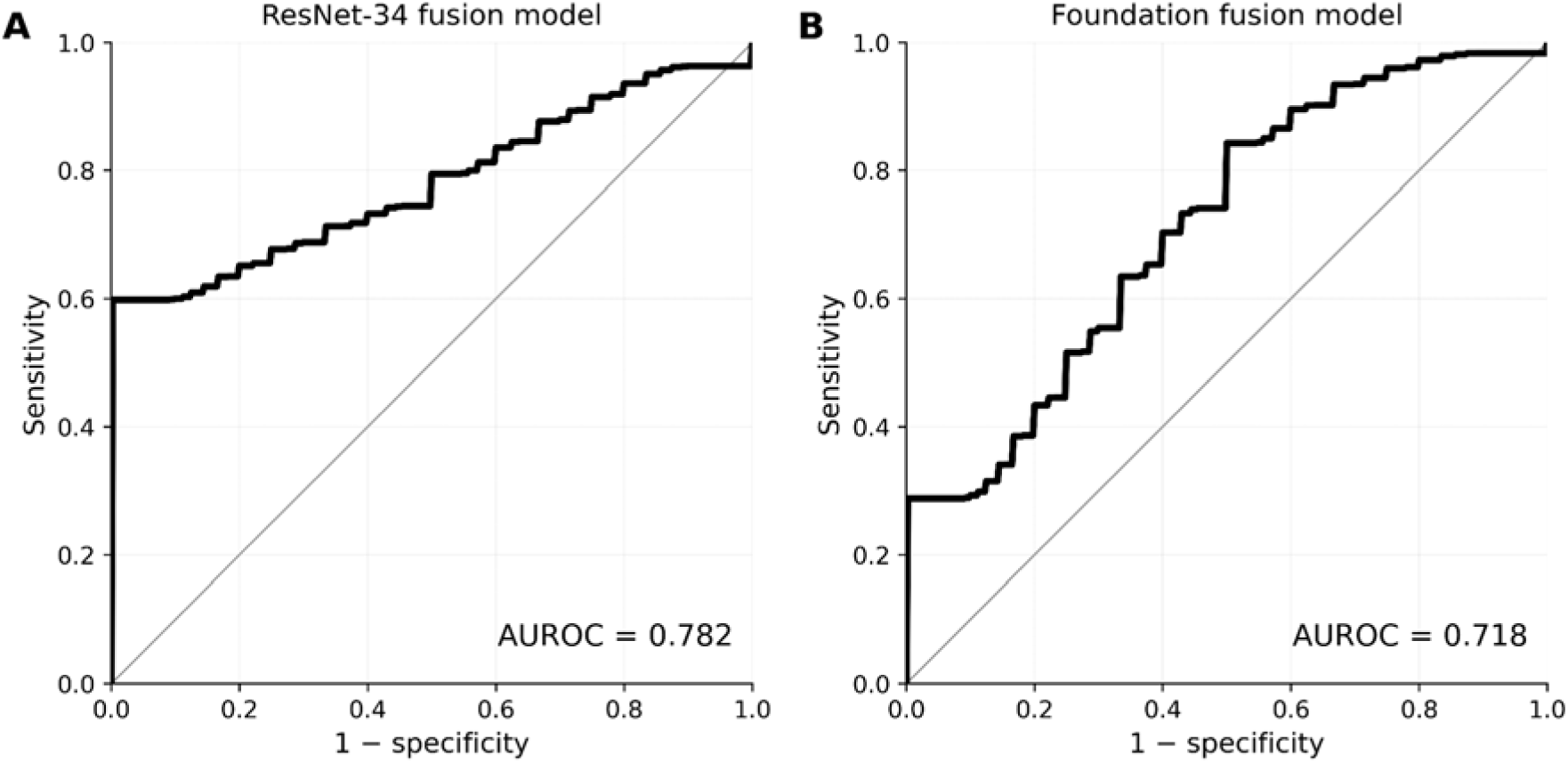
Receiver operating characteristic (ROC) curves for external validation of the multimodal fusion models. Performance on an independent cohort of 19 patients (13 responders, 6 non-responders) from University Hospital Halle. (A) Convolutional branch (ResNet-34) with late fusion, using XGBoost. (B) Foundation model branch with late fusion, using XGBoost. All model components were applied frozen; no retraining, refitting, or recalibration was performed on the external cohort. Area-under-the-receiver-operating-characteristic (AUROC) is shown at the bottom of each panel.

## Discussion

We demonstrated that outcome prediction to ven-aza can be augmented by AI-guided image-based response classification, complementing established gene-based markers of response. In our analysis, multimodal fusion models combining bone marrow smear image data with clinical and genetic information outperformed AI models using either modality alone. For both models - our custom-built ResNet-34-based fusion model and the fine-tuned foundation models - image-based response classification was significantly associated with high prognostic effect sizes for CR achievement in univariable logistic regression, ranking second only to *IDH1/2* mutation status and exceeding those of other genetic alterations including *NPM1*. This effect prevailed in multivariable analysis adjusting for genetic alterations significantly associated with response, and was confirmed via SHAP explainability. Qualitative image-based explainability showed cell-specific model attention, providing a proof-of-concept that model attention is dedicated to cellular composition and morphology, rather than unspecific background signals. In external validation, on a cohort from another center using another WSI scanner and differing staining protocol, our multimodal models still provided accurate predictions.

While risk assessment for AML patients treated with intensive induction chemotherapy is well established, multiple frameworks for patients treated with less-intensive therapies are currently emerging. Given the subpar classification performance of the ELN2022 system^42^ in patients treated with hypomethylating agents and ven^52^, the ELN panel has recently introduced the ELN2024 less-intensive stratification system^7^, including alterations of *IDH1/2, NPM1, DDX41, TP53* and myelodysplasia-related gene mutations. Lachowiez et al.^8^ subsequently introduced the PRISM-3 risk model drawing from 2092 newly diagnosed AML patients treated with HMA and ven, including alterations of *CEBPA, NPM1, IDH1/2, KRAS, JAK2, RUNX1, ASXL1, STAG2*, *BCOR, TP53, DDX41, NRAS, FLT3*-ITD, age, sex, cytogenetics and antecedent disease history, resulting in three risk tiers (low, moderate, high). The authors report better discrimination between risk tiers than with the previously suggested four gene classifier^52^ the ELN2024 system was built upon. Islam et al.^53^ used counterfactual machine learning in newly diagnosed ven-aza treated patients, identifying three tiers (favorable, intermediate, adverse) differentiated by cytogenetics as well as alterations of *IDH1/2, NPM1, BCOR, DNMT3A, DDX41, KRAS, PTPN11, SMC1A, JAK2, KMT2A* and *TP53*. For relapsed and refractory patients, Shahswar et al.^54^ proposed venetoclax prognostic risk score (VEN-PRS) developed using retrospective data from 240 HMA-ven treated patients, including HMA pretreatment, alterations of *NF1, PTPN11, FLT3*, and *TP53* as well as extramedullary disease as predictors of unfavorable outcomes. Importantly, all these risk stratification systems are optimized for survival time and none include image data. In our analysis, prediction of best response was strongly influenced by many of the same genetic alterations identified in these previous studies (most prominently alterations of *IDH1/2*), yet image-based prediction of response outperformed multiple other prognostic variables, including alterations of *NPM1*. Hence, image-based classification could potentially complement existing stratification systems. Adding best response prediction to existing survival time stratification would enable more nuanced decision-making, especially given the ongoing discussion about the role of ven-aza in patients otherwise eligible for intensive induction.^55^

There are several limitations to our study. First, our findings are retrospective and still require prospective validation before clinical implementation. Second, we trained both fusion models on a single center data set using only 193 AML patients and validated on only one external cohort. While good external validation performance using different staining protocols and imaging systems suggests the potential for generalizability, broadening both training and validation cohort across more centers and imaging systems is warranted before claiming full generalizability. Third, our multimodal models encompass imaging, clinical variables (age, blast count) as well as standard NGS-based and cytogenetic markers. Arguably, the effect of adding further modalities such as text-based health records, flow cytometry, histology of core biopsies and inclusion of broader NGS panels or even *omics* data on model performance could be explored. Fourth, we observed a few disparities in outcome predictions between both models: For nine patients, the fusion foundation model correctly predicted CR, while the ResNet-based model was incorrect, and four cases vice versa. While no prognostic stratification model is perfect, this suggests different signals being weighed differently by both model architectures. The marginal differences in performance between both models (foundation model AUROC 0.82; ResNet-based model AUROC 0.79) do not allow us to conclusively prefer one architecture over the other in our analysis yet and therefore warrant a comparative analysis across a broader set of training and validation data. Fifth, extending the inclusion criteria of our models to patients receiving ven-aza in the relapsed/refractory setting and to combination therapies, e.g. in the context of menin inhibition, seems prudent to broaden clinical applicability.

In summary, we developed, fine-tuned, and externally validated two accurate multimodal fusion models incorporating clinical, genetic and bone marrow smear imaging data to predict CR in newly diagnosed AML patients treated with ven-aza. Quantitative and qualitative explainability placed image-based classification among the top prognostic variables and external validation confirmed model generalizability.

## Data Availability

All data are available upon reasonable request to the corresponding author.

## Acknowledgements

This study was supported in part by Deutsche Krebshilfe (German Cancer Aid, Project Number 70117128; granted to J-NE). The funders had no role in conceptualization, design, data collection, analysis, decision to publish, or preparation of the manuscript. The authors gratefully acknowledge the computing time made available to them on the high-performance computer at the NHR Center of TU Dresden. This center is jointly supported by the Federal Ministry of Research, Technology and Space of Germany and the state governments participating in the NHR. Chethan Babu V. Reddy is acknowledged for providing technical support in model deployment.

## Author Contributions

J-NE, IS, JMM, and DAP conceptualized the project. KG, AAO, ZP, MH and DAP provided patient samples. J-NE, KG, AAO, ZP, MMKS, FS, SW, MH, JMM, and DAP annotated patient samples. J-NE and IS developed methodology and implemented deep learning models. All authors analyzed and interpreted the data. J-NE and IS wrote the initial draft. All authors edited the draft, approved the final version of the manuscript, and agreed to be accountable for all aspects of the work.

## Competing Interests

J NE declares consulting services for AstraZeneca, Novartis, and Johnson & Johnson, holds shares in Cancilico, has received an institutional research grant by Novartis, and has received honoraria by Amgen, Astellas, AstraZeneca, Johnson & Johnson, Novartis, Pfizer and Servier. JMM declares consulting services for Johnson & Johnson, Roche, Gilead, Abbvie, Jazz, Pfizer, Astellas, Novartis, AstraZeneca, Clycostem, holds shares in Cancilico and Synagen, has received institutional research grants by Johnson & Johnson, Jazz and Novartis; and has received honoraria by Novartis, Roche, Johnson & Johnson, Abbvie, Pfizer, Sanofi, Astellas and Beigene. MH declares honoraria from Astellas, Daiichi Sankyo, Otsuka, Pfizer, and Servier, paid consultancy for Abbvie, Ascentage Pharma, Astellas, Daiichi Sankyo, Janssen, Miltenyi, and Servier, and research funding to his institution from Abbvie, Bayer Pharma AG, Jazz Pharmaceuticals, Glycostem, PinotBio, Servier, and Toray. The other authors declare no conflict of interest.

## Notes

### Author Declarations

Training and internal testing data was deemed exempt from institutional review board (IRB) review by the IRB of UC Denver, Colorado, USA. Retrospective analysis for external validation data coming from the German Ven/Aza registry was approved by the local Ethics Review Committee (ethical vote No.7972_BO_K_2018), for the Hannover Medical School, Hannover, Germany.

