## Supplemental Information for "Multimodal Artificial Intelligence Fusion Models to Predict Response to Venetoclax and Azacitidine in Patients with Newly Diagnosed Acute Myeloid Leukemia"

### Supplemental Figures

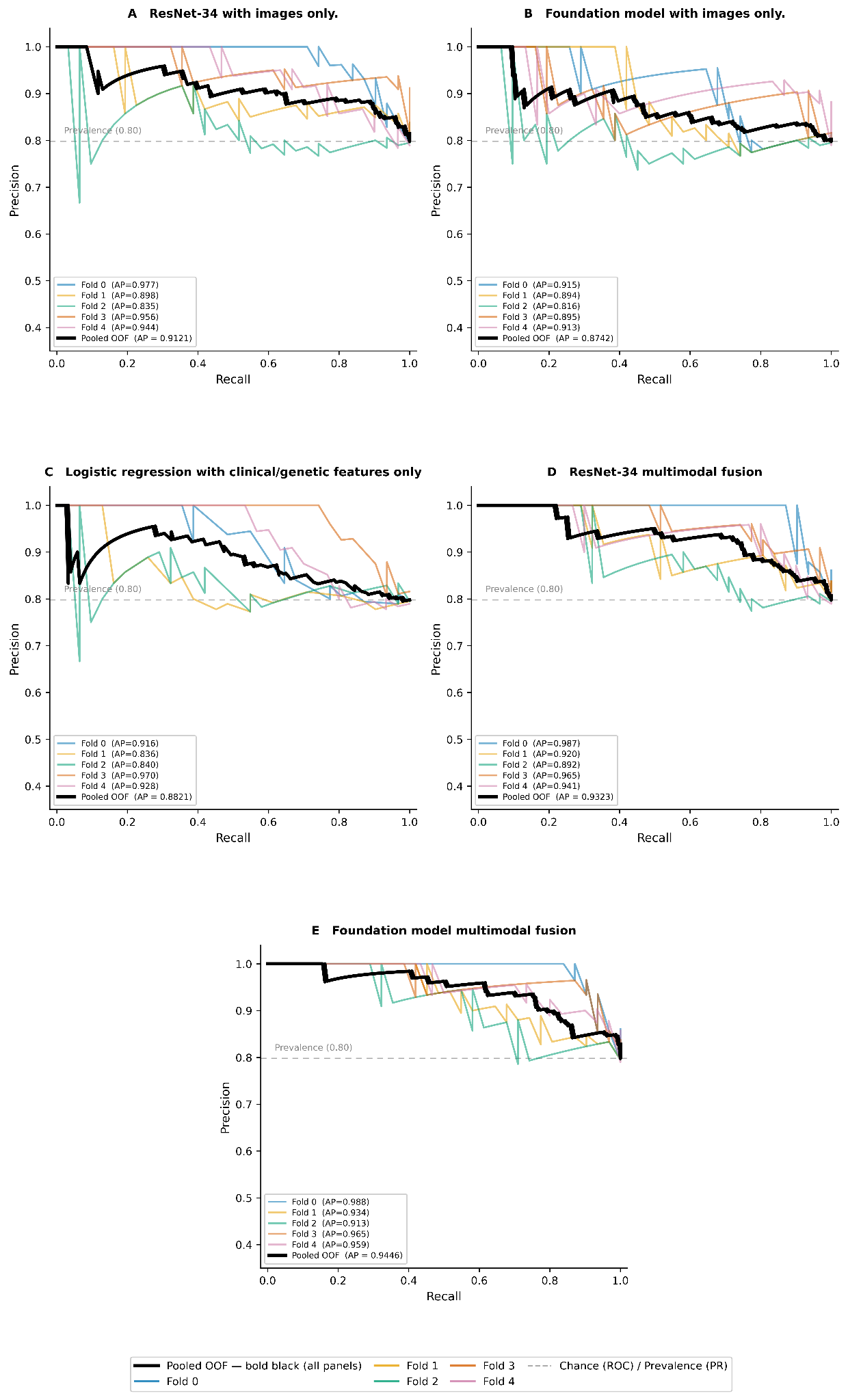

**Supplemental Figure S1. Precision-recall (PR) curves for fusion and non-fusion models evaluated across 5-fold cross-validation.** (A) Image-only custom model (ResNet-34). (B) Image-only foundation model. (C) Clinical/genetic feature-only logistic regression-based classifier. (D) ResNet-34-based fusion model using logistic regression. (E) Foundation model fusion using logistic regression. Individual fold curves are shown in colour; the bold black line represents the pooled out-of-fold (OOF) estimate.

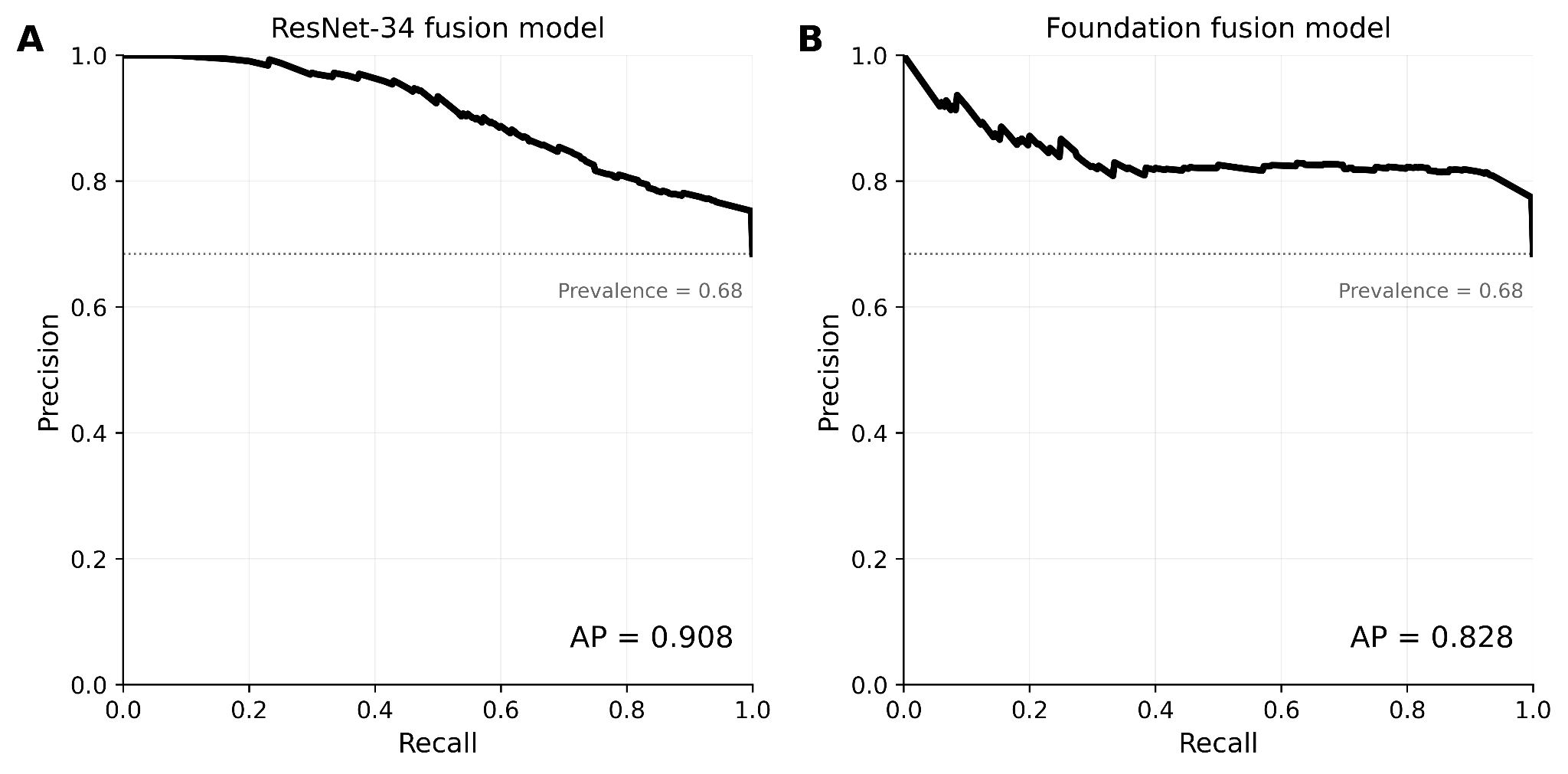

**Supplemental Figure S2. Precision–recall (PR) curves for external validation of the multimodal fusion models.** Performance on an independent cohort of 19 patients (13 responders, 6 non-responders) from University Hospital Halle. (A) Convolutional branch (ResNet-34) with late fusion, using XGBoost. (B) Foundation model branch with late fusion, using XGBoost. Average precision (AP) is shown at the bottom of each panel.

### Supplemental Tables

**Supplemental Table S1. AML patient characteristics**

| **Parameter** |  |
| --- | --- |
| **N** | 193 |
| **Median age, years (IQR)** | 70 (65-76) |
| **Sex, n/N (%)** |  |
| Male | 112/193 (58%) |
| Female | 81/193 (42%) |
| **Bone marrow blast count, % (IQR)** | 50 (30-77) |
| **Molecular genetics, n/N (%)** |  |
| *ASXL1* | 49/193 (25.4%) |
| *FLT3*-ITD | 32/193 (16.6%) |
| *IDH1/2* | 47/193 (24.4%) |
| *NPM1* | 42/193 (21.8%) |
| *PTPN11* | 13/193 (6.7%) |
| *RAS* | 22/193 (11.4%) |
| *RUNX1* | 45/193 (23.3%) |
| *TP53* | 21/193 (10.9%) |
| **ELN2024 less-intensive, n/N (%)**  favorable | 122/193 (63.2%) |
| intermediate | 50/193 (25.9%) |
| adverse | 21/193 (10.9%) |
| **SWOG, n/N (%)** |  |
| favorable | 3/193 (1.6%) |
| intermediate | 93/193 (48.2%) |
| unfavorable / adverse | 69/193 (35.8%) |
| unknown | 28/193 (14.5%) |
| **Response** |  |
| CR/CRi, n/N (%) | 154/193 (79.8%) |

*CR/CRi* complete remission (CR) or complete remission with incomplete hematologic recovery (CRi) in accordance with ELN2022 recommendations, *IQR* interquartile range, *N* number of patients, *SWOG* Southwest Oncology Group/Eastern Cooperative Oncology Group Study system: four cytogenetic risk categories; the unknown category includes cytogenetic aberrations with uncertain prognostic significance

**Supplemental Table S2. Performance metrics for image-only custom CNN (ResNet-34) model in a stratified 5-fold cross-validation set.**

| **Split** | **N** | **Youden Threshold** | **AUROC** | **Accuracy** | **F1** | **Sensitivity** | **Specificity** | **PPV** | **NPV** |
| --- | --- | --- | --- | --- | --- | --- | --- | --- | --- |
| **Fold 0** | 39 | 0.9592 | 0.9052 | 0.7949 | 0.8519 | 0.7419 | 1.0000 | 1.0000 | 0.5000 |
| **Fold 1** | 39 | 0.8437 | 0.7097 | 0.8205 | 0.8889 | 0.9032 | 0.5000 | 0.8750 | 0.5714 |
| **Fold 2** | 39 | 0.9915 | 0.5403 | 0.4872 | 0.5455 | 0.3871 | 0.8750 | 0.9231 | 0.2692 |
| **Fold 3** | 38 | 0.8772 | 0.8525 | 0.9211 | 0.9524 | 0.9677 | 0.7143 | 0.9375 | 0.8333 |
| **Fold 4** | 38 | 0.9213 | 0.8000 | 0.7105 | 0.7843 | 0.6667 | 0.8750 | 0.9524 | 0.4118 |
| **Pooled** | 193 | 0.8437 | 0.7534 | 0.8187 | 0.8867 | 0.8896 | 0.5385 | 0.8839 | 0.5526 |

*AUROC* area-under-the-receiver-operating-characteristic, *CNN* Convolutional Neural Network, *NPV* negative predictive value, *PPV* positive predictive value

**Supplemental Table S3. Performance metrics for image-only UNI+CONCH foundation model in a stratified 5-fold cross-validation set.**

| **Split** | **N** | **Youden Threshold** | **AUROC** | **Accuracy** | **F1** | **Sensitivity** | **Specificity** | **PPV** | **NPV** |
| --- | --- | --- | --- | --- | --- | --- | --- | --- | --- |
| **Fold 0** | 39 | 0.590 | 0.7016 | 0.718 | 0.792 | 0.677 | 0.875 | 0.955 | 0.412 |
| **Fold 1** | 39 | 0.654 | 0.6210 | 0.538 | 0.591 | 0.419 | 1.000 | 1.000 | 0.308 |
| **Fold 2** | 39 | 0.760 | 0.4758 | 0.462 | 0.533 | 0.387 | 0.750 | 0.857 | 0.240 |
| **Fold 3** | 38 | 0.340 | 0.6682 | 0.868 | 0.921 | 0.935 | 0.571 | 0.906 | 0.667 |
| **Fold 4** | 38 | 0.431 | 0.7917 | 0.842 | 0.897 | 0.867 | 0.750 | 0.929 | 0.600 |
| **Pooled** | 193 | 0.676 | 0.6415 | 0.513 | 0.591 | 0.442 | 0.795 | 0.895 | 0.265 |

*AUROC* area-under-the-receiver-operating-characteristic, *NPV* negative predictive value, *PPV* positive predictive value

**Supplemental Table S4. Performance metrics for clinical/genetic data-only logistic regression-based classifier in a stratified 5-fold cross-validation set.**

| **Split** | **N** | **Youden Threshold** | **AUROC** | **Accuracy** | **F1** | **Sensitivity** | **Specificity** | **PPV** | **NPV** |
| --- | --- | --- | --- | --- | --- | --- | --- | --- | --- |
| **Fold 0** | 39 | 0.7883 | 0.6996 | 0.615 | 0.703 | 0.548 | 0.875 | 0.944 | 0.333 |
| **Fold 1** | 39 | 0.8034 | 0.5323 | 0.385 | 0.400 | 0.258 | 0.875 | 0.889 | 0.233 |
| **Fold 2** | 39 | 0.6306 | 0.5726 | 0.821 | 0.896 | 0.968 | 0.250 | 0.833 | 0.667 |
| **Fold 3** | 38 | 0.7730 | 0.8710 | 0.789 | 0.852 | 0.742 | 1.000 | 1.000 | 0.467 |
| **Fold 4** | 38 | 0.7692 | 0.7396 | 0.632 | 0.696 | 0.533 | 1.000 | 1.000 | 0.364 |
| **Pooled** | 193 | 0.7882 | 0.6643 | 0.539 | 0.619 | 0.461 | 0.846 | 0.922 | 0.284 |

*AUROC* area-under-the-receiver-operating-characteristic, *NPV* negative predictive value, *PPV* positive predictive value

**Supplemental Table S5. Performance metrics for UNI+CONCH foundation model with late fusion using logistic regression in a stratified 5-fold cross-validation set.**

| **Split** | **N** | **Youden Threshold** | **AUROC** | **Accuracy** | **F1** | **Sensitivity** | **Specificity** | **PPV** | **NPV** |
| --- | --- | --- | --- | --- | --- | --- | --- | --- | --- |
| **Fold 0** | 39 | 0.486 | 0.9476 | 0.897 | 0.9310 | 0.871 | 1.000 | 1.000 | 0.667 |
| **Fold 1** | 39 | 0.798 | 0.7661 | 0.565 | 0.622 | 0.452 | 1.000 | 1.000 | 0.320 |
| **Fold 2** | 39 | 0.623 | 0.7016 | 0.641 | 0.720 | 0.581 | 0.875 | 0.947 | 0.350 |
| **Fold 3** | 38 | 0.429 | 0.8710 | 0.895 | 0.933 | 0.903 | 0.857 | 0.966 | 0.667 |
| **Fold 4** | 38 | 0.454 | 0.8625 | 0.763 | 0.830 | 0.733 | 0.875 | 0.957 | 0.467 |
| **Pooled** | 193 | 0.504 | 0.8170 | 0.761 | 0.835 | 0.753 | 0.795 | 0.935 | 0.449 |

*AUROC* area-under-the-receiver-operating-characteristic, *NPV* negative predictive value, *PPV* positive predictive value

**Supplemental Table S6. Performance metrics for UNI+CONCH foundation model with late fusion using ElasticNet in a stratified 5-fold cross-validation set.**

| **Split** | **N** | **Youden Threshold** | **AUROC** | **Accuracy** | **F1** | **Sensitivity** | **Specificity** | **PPV** | **NPV** |
| --- | --- | --- | --- | --- | --- | --- | --- | --- | --- |
| **Fold 0** | 39 | 0.459 | 0.8952 | 0.846 | 0.900 | 0.839 | 0.875 | 0.966 | 0.538 |
| **Fold 1** | 39 | 0.553 | 0.7823 | 0.718 | 0.792 | 0.677 | 0.875 | 0.955 | 0.412 |
| **Fold 2** | 39 | 0.527 | 0.6815 | 0.641 | 0.720 | 0.581 | 0.875 | 0.947 | 0.350 |
| **Fold 3** | 38 | 0.508 | 0.9234 | 0.816 | 0.865 | 0.774 | 1.000 | 1.000 | 0.500 |
| **Fold 4** | 38 | 0.396 | 0.7854 | 0.711 | 0.774 | 0.667 | 0.875 | 0.909 | 0.467 |
| **Pooled** | 193 | 0.545 | 0.8067 | 0.741 | 0.810 | 0.695 | 0.923 | 0.958 | 0.470 |

*AUROC* area-under-the-receiver-operating-characteristic, *NPV* negative predictive value, *PPV* positive predictive value

**Supplemental Table S7. Performance metrics for UNI+CONCH foundation model with late fusion using random forest in a stratified 5-fold cross-validation set.**

| **Split** | **N** | **Youden Threshold** | **AUROC** | **Accuracy** | **F1** | **Sensitivity** | **Specificity** | **PPV** | **NPV** |
| --- | --- | --- | --- | --- | --- | --- | --- | --- | --- |
| **Fold 0** | 39 | 0.580 | 0.8589 | 0.846 | 0.897 | 0.839 | 0.875 | 0.966 | 0.538 |
| **Fold 1** | 39 | 0.560 | 0.7177 | 0.692 | 0.769 | 0.645 | 0.875 | 0.952 | 0.389 |
| **Fold 2** | 39 | 0.580 | 0.6633 | 0.590 | 0.667 | 0.548 | 0.750 | 0.895 | 0.310 |
| **Fold 3** | 38 | 0.520 | 0.9437 | 0.842 | 0.897 | 0.806 | 1.000 | 1.000 | 0.538 |
| **Fold 4** | 38 | 0.400 | 0.7708 | 0.684 | 0.741 | 0.633 | 0.875 | 0.905 | 0.438 |
| **Pooled** | 193 | 0.612 | 0.7913 | 0.725 | 0.790 | 0.675 | 0.923 | 0.946 | 0.448 |

*AUROC* area-under-the-receiver-operating-characteristic, *NPV* negative predictive value, *PPV* positive predictive value

**Supplemental Table S8. Performance metrics for UNI+CONCH foundation model with late fusion using XGBoost in a stratified 5-fold cross-validation set.**

| **Split** | **N** | **Youden Threshold** | **AUROC** | **Accuracy** | **F1** | **Sensitivity** | **Specificity** | **PPV** | **NPV** |
| --- | --- | --- | --- | --- | --- | --- | --- | --- | --- |
| **Fold 0** | 39 | 0.903 | 0.8790 | 0.872 | 0.912 | 0.839 | 1.000 | 1.000 | 0.615 |
| **Fold 1** | 39 | 0.981 | 0.7177 | 0.667 | 0.750 | 0.613 | 0.875 | 0.952 | 0.412 |
| **Fold 2** | 39 | 0.978 | 0.7258 | 0.769 | 0.842 | 0.742 | 0.875 | 0.958 | 0.438 |
| **Fold 3** | 38 | 0.992 | 0.9585 | 0.895 | 0.931 | 0.871 | 1.000 | 1.000 | 0.636 |
| **Fold 4** | 38 | 0.940 | 0.7396 | 0.553 | 0.645 | 0.467 | 0.875 | 1.000 | 0.350 |
| **Pooled** | 193 | 0.966 | 0.7752 | 0.756 | 0.822 | 0.714 | 0.923 | 0.964 | 0.463 |

*AUROC* area-under-the-receiver-operating-characteristic, *NPV* negative predictive value, *PPV* positive predictive value

**Supplemental Table S9. Performance metrics for CNN (ResNet-34) architecture with late fusion using logistic regression in a stratified 5-fold cross-validation set.**

| **Split** | **N** | **Youden Threshold** | **AUROC** | **Accuracy** | **F1** | **Sensitivity** | **Specificity** | **PPV** | **NPV** |
| --- | --- | --- | --- | --- | --- | --- | --- | --- | --- |
| **Fold 0** | 39 | 0.524 | 0.9435 | 0.923 | 0.949 | 0.903 | 1.000 | 1.000 | 0.727 |
| **Fold 1** | 39 | 0.479 | 0.7419 | 0.795 | 0.867 | 0.839 | 0.625 | 0.897 | 0.500 |
| **Fold 2** | 39 | 0.491 | 0.6371 | 0.615 | 0.706 | 0.581 | 0.750 | 0.900 | 0.316 |
| **Fold 3** | 38 | 0.577 | 0.8571 | 0.789 | 0.857 | 0.7741 | 0.857 | 0.960. | 0.462 |
| **Fold 4** | 38 | 0.362 | 0.8167 | 0.816 | 0.873 | 0.800 | 0.875 | 0.960 | 0.538 |
| **Pooled** | 193 | 0.519 | 0.7886 | 0.736 | 0.813 | 0.721 | 0.795 | 0.933 | 0.419 |

*AUROC* area-under-the-receiver-operating-characteristic, *CNN* Convolutional Neural Network, *NPV* negative predictive value, *PPV* positive predictive value

**Supplemental Table S10. Confusion matrices for the foundation fusion model and custom CNN (ResNet-34) architecture, both with late fusion using logistic regression.**

| **Foundation model fusion** | | | **Custom CNN model fusion** | | |
| --- | --- | --- | --- | --- | --- |
| **GT/Pred** | **Pred resp.** | **Pred refr.** | **GT/Pred** | **Pred resp.** | **Pred refr.** |
| **GT resp.** | 116  TP (60.1%) | 38  FN (19.7%) | **GT resp.** | 111  TP (57.5%) | 43  FN (22.3%) |
| **GT refr.** | 8  FP (4.1%) | 31  TN (16.1%) | **GT refr.** | 9  FP (4.7%) | 30  TN (15.5%) |

*CNN* Convolutional Neural Network, *FN* false negative, *FP* false positive, *GT* Ground Truth, *Pred* Prediction, *refr.* Refractory, *resp.* Responder, *TN* true negative, *TP* true positive

**Supplemental Table S11. Detailed information of the discordant cases between foundation model and custom CNN (ResNet-34) fusion models.**

| **ID** | **GT** | **FM Pred** | **ResNet Pred** | **ELN 2024** | **Age** | **Molecular** |
| --- | --- | --- | --- | --- | --- | --- |
| 0 | CR/CRi | TP | FN | Adverse | 71-75 | *TP53* |
| 1 | CR/CRi | TP | FN | Adverse | 66-70 | *TP53* |
| 2 | CR/CRi | TP | FN | Adverse | 80-85 | *TP53* |
| 3 | CR/CRi | TP | FN | Adverse | 66-70 | *TP53*, *GATA2* |
| 4 | CR/CRi | TP | FN | Adverse | 66-70 | *TP53* |
| 5 | CR/CRi | TP | FN | Intermediate | 66-70 | *BCOR*, *KRAS*, *PTPN11*, *RUNX1*, *U2AF1* |
| 6 | CR/CRi | TP | FN | Favorable | 81-85 | *CUX1*, *PTPN11*, *RUNX1*, *TET2*; *EVI1* rearrangement |
| 7 | CR/CRi | TP | FN | Intermediate | 36-40 | *IDH2*, *ASXL1*, *NRAS*, *PTPN11*, *SMC1A* |
| 8 | CR/CRi | TP | FN | Favorable | 66-70 | *DDX41* |
| 9 | CR/CRi | FN | TP | Adverse | 66-70 | *TP53* |
| 10 | CR/CRi | FN | TP | Adverse | 76-80 | *TP53*, *DNMT3A*, *BCOR* |
| 11 | CR/CRi | FN | TP | Favorable | 71-75 | *ASXL1*, *KMT2A*, *SETBP1*, *PHF6* |
| 12 | CR/CRi | FN | TP | Favorable | 56-60 | *DNMT3A*, *FLT3*-TKD, *CEBPA*, *RUNX1*, *U2AF1*, *BCOR*, *BCORL1* |
| 13 | Refractory | TN | FP | Intermediate | 66-70 | *FLT3*-ITD, *RUNX1*, *TET2*, *MPL* |
| 14 | Refractory | TN | FP | Adverse | 51-55 | *TP53* |
| 15 | Refractory | FP | TN | Adverse | 71-75 | *TP53*, *GATA2* |
| 16 | Refractory | FP | TN | Favorable | 66-70 | *ASXL1*, *RUNX1*, *TET2* |

*CR/CRi* complete remission (CR) or complete remission with incomplete hematologic recovery (CRi) in accordance with ELN2022 recommendations, *FM* foundation model, *FN* false negative, *FP* false positive, *Pred* Prediction, *TN* true negative, *TP* true positive

**Supplemental Table S12. Performance metrics of four heads for the CNN (ResNet-34) fusion model validated on the external dataset.**

| **Head** | | **Youden Threshold** | **AUROC across folds**  **(95%-CI)** | **Accuracy** | **F1** | **Sensitivity** | **Specificity** | **PPV** | **NPV** |
| --- | --- | --- | --- | --- | --- | --- | --- | --- | --- |
| LR | | 0.519 | 0.636 ± 0.023 (0.33–0.91) | 0.632 (0.41–0.81) | 0.720 (0.48–0.89) | 0.692 (0.42–0.87) | 0.500 (0.19–0.81) | 0.750 (0.47–0.91) | 0.429 (0.16–0.75) |
| Elasticnet | | 0.597 | 0.681 ± 0.031 (0.37–0.92) | 0.632 (0.41–0.81) | 0.720 (0.48–0.89) | 0.692 (0.42–0.87) | 0.500 (0.19–0.81) | 0.750 (0.47–0.91) | 0.429 (0.16–0.75) |
| RF | | 0.635 | 0.715 ± 0.055 (0.37–1.00) | 0.684 (0.46–0.85) | 0.769 (0.55–0.92) | 0.769 (0.50–0.92) | 0.500 (0.19–0.81) | 0.769 (0.50–0.92) | 0.500 (0.19–0.81) |
| XGBoost | | 0.834 | 0.769 ± 0.055 (0.54–0.97) | 0.737 (0.51–0.88) | 0.828 (0.64–0.95) | 0.923 (0.67–0.99) | 0.333 (0.10–0.70) | 0.750 (0.51–0.90) | 0.667 (0.21–0.94) |

*AUROC* area-under-the-receiver-operating-characteristic, *LR* logistic regression, *NPV* negative predictive value, *PPV* positive predictive value, *RF* random forest

**Supplemental Table S13. Performance metrics of four heads for the foundation model fusion validated on the external dataset.**

| **Head** | | **Youden Threshold** | **AUROC**  **across folds**  **(95%-CI)** | **Accuracy** | **F1** | **Sensitivity** | **Specificity** | **PPV** | **NPV** |
| --- | --- | --- | --- | --- | --- | --- | --- | --- | --- |
| LR | | 0.504 | 0.600 ± 0.025 (0.32-0.89) | 0.684 (0.46–0.85) | 0.769 (0.55–0.92) | 0.769 (0.50–0.92) | 0.500 (0.19–0.81) | 0.769 (0.50–0.92) | 0.500 (0.19–0.81) |
| Elasticnet | | 0.545 | 0.651 ± 0.019  (0.33–0.92) | 0.684 (0.46–0.85) | 0.769 (0.55–0.92) | 0.769 (0.50–0.92) | 0.500 (0.19–0.81) | 0.769 (0.50–0.92) | 0.500 (0.19–0.81) |
| RF | | 0.612 | 0.636 ± 0.039 (0.27–1.00) | 0.684 (0.46–0.85) | 0.769 (0.55–0.92) | 0.769 (0.50–0.92) | 0.500 (0.19–0.81) | 0.769 (0.50–0.92) | 0.500 (0.19–0.81) |
| XGBoost | | 0.966 | 0.733 ± 0.026 (0.36–1.00) | 0.684 (0.46–0.85) | 0.769 (0.55–0.92) | 0.769 (0.50–0.92) | 0.500 (0.19–0.81) | 0.769 (0.50–0.92) | 0.500 (0.19–0.81) |

*AUROC* area-under-the-receiver-operating-characteristic, *LR* logistic regression, *NPV* negative predictive value, *PPV* positive predictive value, *RF* random forest

**Supplemental Table S14. Confusion matrices for the foundation fusion model and custom CNN (ResNet-34) fusion model, both with late fusion using XGBoost, validated on the external dataset.**

| **Foundation model fusion** | | | **Custom CNN model fusion** | | |
| --- | --- | --- | --- | --- | --- |
| **GT/Pred** | **Pred resp.** | **Pred refr.** | **GT/Pred** | **Pred resp.** | **Pred refr.** |
| **GT resp.** | 10  TP (52.6%) | 3  FN (15.8%) | **GT resp.** | 12  TP (63.2%) | 1  FN (5.3%) |
| **GT refr.** | 3  FP (15.8%) | 3  TN (15.8%) | **GT refr.** | 4  FP (21.1%) | 2  TN (10.5%) |

*CNN* Convolutional Neural Network, *FN* false negative, *FP* false positive, *GT* Ground Truth, *Pred* Prediction, *refr.* Refractory, *resp.* Responder, *TN* true negative, *TP* true positive
